# A stepwise short- and long-read whole genome sequencing strategy resolves previous genetically unresolved inherited retinal disease cases

**DOI:** 10.64898/2026.09.23.26363559

**Authors:** Kim Rodenburg, Stefanida Shliaga, Lonneke Haer-Wigman, Anneke T. Vulto-van Silfhout, Erica G.M. Boonen, Lara K. Holtes, Galuh D.N. Astuti, Wolfgang Berger, Tamar Ben-Yosef, Camiel J.F. Boon, Ronny Derks, G. Jane Farrar, Christian Gilissen, Elise Héon, Carel B. Hoyng, Caroline C.W. Klaver, Hester Y. Kroes, Petra Liskova, Monika Ołdak, Dominika Oziębło, Jacoline B. ten Brink, Astrid S. Plomp, Alberta A.H.J. Thiadens, Joke Verheij, Marianna Weener, Susanne Kohl, Frans P.M. Cremers, L. Ingeborgh van den Born, Suzanne E. de Bruijn, Susanne Roosing

**Affiliations:** Department of Human Genetics, Radboud University Medical Center, Geert Grooteplein Zuid 10, Nijmegen, The Netherlands; Department of Clinical Genetics, Maastricht University Medical Center, Maastricht, The Netherlands; The Rotterdam Eye Hospital, Rotterdam Ophthalmic Institute, Rotterdam, The Netherlands; Institute of Medical Molecular Genetics, University of Zurich, Schlieren, Switzerland; Neuroscience Center Zurich, University and ETH Zurich, Zurich, Switzerland; Center for Integrative Human Physiology, University of Zurich, Zurich, Switzerland; Ruth and Bruce Rappaport Faculty of Medicine, Technion-Israel Institute of Technology, Haifa, Israel; Department of Ophthalmology, Leiden University Medical Center, Leiden, The Netherlands; Department of Ophthalmology, Amsterdam University Medical Center, Amsterdam, The Netherlands; Institute of Genetics, School of Genetics and Microbiology, Trinity College Dublin, The University of Dublin; Department of Ophthalmology and Vision Sciences, Ocular Genetics Program, The Hospital for Sick Children, 555 University Avenue, Toronto, Ontario, Canada; Department of Ophthalmology, Radboud University Medical Center, Geert Grooteplein Zuid 10, Nijmegen, The Netherlands; Department of Ophthalmology, Erasmus MC University Medical Center, Rotterdam, The Netherlands; Division Laboratories, Pharmacy and Biomedical Genetics, Department of Genetics, University Medical Center of Utrecht, Utrecht, The Netherlands; Department of Paediatrics and Inherited Metabolic Disorders, First Faculty of Medicine, Charles University and General University Hospital in Prague, Prague, Czech Republic; Department of Ophthalmology, First Faculty of Medicine, Charles University and General University Hospital in Prague, Prague, Czech Republic; Department of Histology and Embryology, Medical University of Warsaw, T. Chałubińskiego 5, Warsaw, Poland; Department of Genetics, Institute of Physiology and Pathology of Hearing, M. Mochnackiego 10, Warsaw, Poland; Department of Human Genetics, Amsterdam University Medical Center, University of Amsterdam, Amsterdam, The Netherlands; Department of Medical Genetics, University Medical Center Groningen, University of Groningen, Groningen, The Netherlands; CRO Oftalmic, Moscow, Russia; Institute for Ophthalmic Research, Centre for Ophthalmology, University Hospital Tübingen, Elfriede-Aulhorn-Str. 7, Tübingen, Germany; Department of Otorhinolaryngology, Hearing & Genes, Radboud University Medical Center, Geert Grooteplein Zuid 10, Nijmegen, The Netherlands

## Abstract

**Purpose:** A significant proportion of cases with rare inherited retinal disease (IRD) remain genetically unresolved following short-read whole genome sequencing (WGS). This study aimed to increase the diagnostic yield of two cohorts in a research setting. One cohort was previously screened by short-read WGS (n=120 cases), while the second cohort (n=28 cases) was not included in any previous short-read WGS studies. In contrast to the larger cohort, these cases only underwent either exome sequencing (ES) or single-molecule molecular inversion probe (smMIPs) sequencing as pre-screening in earlier studies. For all cases, we performed a stepwise, case-by-case reanalysis in which short-read WGS was generated (n=28 cases) and (re)analyzed short-read WGS data using an optimized approach. For a subset of cases (n=20) that remained unresolved, this was followed by long-read WGS. Together, these steps aimed to improve the diagnostic yield in these cohorts.

**Methods:** Short-read WGS data of 148 IRD-cases were (re)analyzed using updated allele frequency databases, new variant caller and variant predictor tools, as well as updated and extended gene-panels to identify causal single nucleotide variants (SNVs) and structural variants (SVs). For 20 genetically unresolved cases with sufficient high-quality DNA available, long-read WGS was performed.

**Results:** The (re)analysis of short-read WGS data in the 120 previously WGS-screened unresolved cases resulted in a diagnostic yield of 15% (18/120). For the 28 individuals without prior WGS, performing WGS analysis within this study provided an additional yield of 32% (9/28). Furthermore, long-read WGS contributed two genetic diagnoses among the 20 long-read WGS-sequenced probands. Amongst other reasons, new genetic diagnoses were attributed to pathogenic variants in genes newly associated with IRDs and in genes that were not included in the gene panels applied in previous WGS studies. The Mobile Element Locator Tool facilitated the identification of two pathogenic Alu insertions in short-read WGS. Long-read WGS identified two additional pathogenic variants.

**Conclusion:** These results support periodic reanalysis of existing short-read WGS data as a key component of IRD genetic diagnostics, with selective long-read WGS representing a valuable additional approach for unresolved cases. Together, these complementary strategies offer a practical framework for improving molecular diagnosis and narrowing the remaining diagnostic gap in IRDs.

## Introduction

Inherited retinal diseases (IRDs) are a diverse group of rare and clinically and genetically heterogeneous disorders, that collectively represent a major cause of visual impairment, with a global prevalence estimate between 1:1,000-1:4,000^1^. IRDs are predominantly monogenic syndromic or non-syndromic diseases and vary in age of onset, course, severity and rate of disease progression, with overlapping clinical features for the specific subtypes of IRDs. Most IRDs follow Mendelian inheritance patterns (autosomal recessive (AR), autosomal dominant (AD), X-linked (XL)), with AR being the most frequent pattern of disease. While rare non-Mendelian inheritance patterns, such as mitochondrial and digenic inheritance patterns have been reported, this merely represents a fraction of all cases^2^. To date, RetNet and RetiGene currently list 348 and 527 IRD-associated genes (RetNet, https://RetNet.org/; RetiGene V1.12, accessed July 2026)^3^.

The general need for a genetic diagnosis has driven advances in sequencing technologies used in genetic testing, including their application for genetic testing in unresolved IRD cases. While targeted approaches, such as Sanger sequencing, targeted panel sequencing and exome sequencing (ES) have been shown to be effective in discovering pathogenic variants in the coding regions of the genome, these methods are now being complemented and expanded by more comprehensive methods such as short-read whole genome sequencing (WGS). Collectively, these approaches result in diagnostic yield varying between 28-64%, depending on the composition of the cohort and technique applied^1,4–12^. Part of this yield reflects the specific contribution of WGS when applied to cases that were negative following pre-screening with targeted approaches. In these cases, WGS has been shown to resolve an additional 9.6-31%^12–15^. The remaining missing heritability in these unresolved IRD cases is likely attributable to several factors. A contributing factor are the technical limitations of short-read WGS. The limited coverage and mapping performance in low-complexity regions, incomplete detection of complex structural variants (SVs), and poor sensitivity for mobile element insertions. However, the missing heritability may as well stem from gaps in current variant and disease-gene knowledge, variants that are technically detectable by short-read WGS but are overlooked, misclassified, or not yet linked to disease due to undiscovered gene-disease associations. Distinguishing between a technical detection problem, which could be addressed through long-read WGS, and a knowledge gap, which requires systematic reanalysis of existing data, is therefore essential, and formed the rationale for first conducting a reanalysis approach prior to long-read WGS.

In this study, four and five years after the initial WGS studies by Fadaie et al. and Reurink et al., we aimed to increase the diagnostic yield in these cohorts of unresolved cases (n=121) of individuals representing the broad spectrum of IRDs through a case-by-case (re)analysis approach in a research setting^14,16^. We additionally included a second cohort of 28 cases that had not undergone prior ES or WGS. First, short-read WGS was generated for the 28 cases which prior WGS data and data were (re)analyzed using a refined variant identification framework, including updated IRD gene and variant databases, novel *in silico* variant callers and prediction tools, and functional assays. Second, a subset (n=20) of cases that remained without a genetic diagnosis underwent long-read WGS to explore previously undetected variants.

## Material and Methods

A total of 148 index IRD cases were included in this study (**Table 1, Supplementary table 1**). Written informed consent was obtained by the corresponding centers, adherent to the tenets of the declaration of Helsinki and as approved by the local ethics committee of the Radboud University Medical Center Nijmegen, as an amendment to the approval by the local ethics committee of the Rotterdam Eye Hospital (MEC-2010-359; OZR protocol no. 2009-32). Ethics board of the Medical Faculty of the University of Tübingen (116/2015BO2). The Royal Victoria Eye and Ear Hospital (Dublin, Ireland) (13-06-2011: HRA-POR201097), Rambam Health Care Campus (Haifa, Israel), Tel Aviv Sourasky Medical Center (Tel Aviv, Israel), Bnai Zion Medical Center (Haifa, Israel), Schneider Children’s Medical Center of Israel (Petach Tikva, Israel) and HaEmek Medical Center (Afula, Israel).

**Table 1:** Overview of all phenotypes present in the cohort (n=148)

| Phenotype | Number of cases | Percentage in cohort |
| --- | --- | --- |
| Retinitis pigmentosa | 80 | 54% |
| Usher syndrome | 19 | 13% |
| Cone-rod dystrophy | 10 | 7% |
| Macular dystrophy | 13 | 9% |
| Stargardt disease | 5 | 3% |
| Rod dystrophy | 5 | 3% |
| Cone dystrophy | 4 | 3% |
| Leber congenital amaurosis | 2 | 1% |
| Achromatopsia | 2 | 1% |
| Retinal pigment epithelium dystrophy | 2 | 1% |
| Rod cone dystrophy | 2 | 1% |
| Bardet-Biedl syndrome | 1 | 1% |
| Congenital stationary night blindness | 1 | 1% |
| Nystagmus | 1 | 1% |
| Fundus albipunctatus | 1 | 1% |
| **Total** | **148** | **100%** |

### Short-read whole genome sequencing

Genomic DNA from probands was isolated from peripheral blood lymphocytes according to standard procedures. WGS was performed at BGI on a BGISeq500 using 2× 150 base pair (bp) paired-end reads with a minimal median coverage of 30-fold per genome. Sequencing reads were mapped to the Human Reference Genome build GRCh38/hg38 using Burrows-Wheeler Aligner V.0.7814^17^. Single nucleotide variants (SNVs) and small indels (<50 bp) were called using Genome Analysis Toolkit HaplotypeCaller (Broad Institute), structural variants (SVs) were called using Manta Structural Variant Caller, based on read-pair evidence and read-depth evidence^18,19^. Copy number variants were called using Canvas Copy Number Variant Caller, based on read-depth evidence, mobile element insertions (MEI) were identified using the Mobile Element Insertion Tool (MELT). SNVs, small indels, SVs and CNVs were annotated using an in-house pipeline^20,21^.

### Long-read whole genome sequencing

Long-read WGS and variant calling was performed for selected cases as previously described^22^. In short, seven micrograms of genomic DNA were sheared to an average size of ∼15-18 kb and library preparation was performed using the SMRTbell Prep kit 3.0 (PacBio) followed by size selection using the BluePippin system. Sequencing primers and polymerase were annealed to the SMRTbell library using the Sequel II binding kit 3.2 (PacBio), before loading on an 8M SMRTcell and sequencing (30 h movie time) on the Sequel IIe system using a single flow cell. HiFi-sequencing reads were generated using the SMRT Link 8.0.0 software (PacBio) and mapped against genome build GRCh38). Assessment of prioritized variants was performed by visual inspection of the sequencing reads in the Integrative Genomics Viewer (IGV, version 2.17) software^23^.

### Variant prioritization and (re)analysis

WGS data were analyzed to identify potential pathogenic variants in all IRD-associated genes as listed on RetNet (https://sph.uth.edu/retnet/, accessed July, 2026) as well as in genes only recently associated with IRD that are not published in RetNet. Coding and non-coding SNVs were selected based on a minor allele frequency of <0.01 in the gnomAD population database (all populations) (version 3.1.2, with additional manual checks in v4.1.1 for selected variants). Next, the selected rare variants were prioritized based on protein effect. Nonsense, start- or stop-affecting variants, frameshift, in-frame, missense variants, canonical and intronic splice variants were examined in more detail. For missense variants, CADD_PHRED and REVEL scores were obtained. Variants meeting the predefined thresholds of both *in silico* prediction tools (CADD_PHRED: >15, range 0-99; REVEL: >0.3, range 0-1) were selected for further investigation as possibly pathogenic candidates^24,25^. Variants meeting the threshold of a single tool were retained as lower-priority candidate variants. Similarly, SpliceAI delta scores (https://spliceailookup.broadinstitute.org/) were obtained for all variants with a potential effect on splicing, including canonical splice variants, missense, synonymous and intronic variants^26^. Variants were selected for extended investigation and potential subsequent further testing if at least one out of the four provided scores (acceptor gain (DS_AG), acceptor loss (DS_AL), donor gain (DS_DG) or donor loss (DS_DL)) provided a delta score of ≥ 0.2 using the following window setting: upstream and downstream 50 bp. Coding SVs, including CNVs, were prioritized on a minor allele frequency of <0.01 in the 1000 genome database^27^. Intronic SVs, including CNVs, were only selected for further evaluation when a monoallelic variant was present in the same gene. Inversions and duplications were considered potentially pathogenic when disrupting an IRD-associated gene, as proposed by de Bruijn et al^28^. We prioritized MEI in the coding, splice and untranslated (UTR)-regions of genes and only considered insertions in the intronic regions of a gene in case a monoallelic variant was identified in the respective gene for that case.

### Aberrant splicing analysis: *in vitro* minigene splice assay

Minigene constructs were generated as previously described^29^. To summarize, at least 450 bp of the variant’s up- and downstream flanking intronic sequences were amplified from the proband’s genomic DNA. Both wildtype fragments and fragments containing the selected variants were cloned into an adapted pCl-NEO vector using Gateway cloning technology (Thermo Fisher Scientific, Carlsbad, CA, USA), between *RHO* exons 3 and 5. HEK293T cells were transfected using polyethylenimine (PEI) with either a wildtype or mutant minigene construct and harvested 48 hours post transfection. RNA was isolated using the Nucleospin RNA kit (Machery-Nagel, Düren, Germany) following manufacturer’s instructions. One microgram of total RNA was used as input for cDNA synthesis using the iScript cDNA synthesis kit (Bio-Rad, Hercules, CA, USA) according to manufacturer’s instructions. The reverse transcription-polymerase chain reactions (RT-PCR) were performed using primers for *RHO* exons 3 and 5 to assess the splicing pattern of the regions of interest. Primers to amplify *ACTB* were used as a loading control. PCR fragments were sequence-verified by Sanger sequencing. Transfections were performed in two independent experiments.

### Aberrant splicing analysis: targeted long-read cDNA transcript analysis

The effect of the *BBS4* variant was confirmed using targeted long-read cDNA transcript analysis as described previously^30^. In short, a fresh blood sample was collected in a PAXgene Blood RNA tube from the proband carrying a *BBS4* c.24+40C>T variant. RNA was isolated using the PAXgene Blood RNA kit (Qiagen, Hilden, Germany) according to the manufacturer’s protocol and RNA quantification was performed by DeNovix (Wilmington, DE). cDNA synthesis was performed using SuperScript IV Reverse Transcriptase (Invitrogen) with random hexamers following an adjusted protocol to enrich for long cDNA molecules. 500 ng input RNA was incubated at 60°C for 10 min to allow for the linearization of long molecules. In addition, cDNA incubation was performed at 55°C for 50 min instead of the recommended 10 min. One microliter undiluted cDNA was used as input for RT-PCR performed using standard PCR conditions and primers designed to amplify from exon 1 to exon 10 of *BBS4*. Targeted long-read cDNA sequencing and transcript analysis was performed as described previously^30^. Sequencing results were analyzed using IGV software version 2.17. The percentage of altered transcripts was calculated using the number of reads spanning specific exon-exon junctions, according to the formula: (number of reads covering a specific exon-exon junction / total number of reads) × 100%.

### Breakpoint analysis

Breakpoints of the identified *PCDH15* SV and *USH2A* MEI were confirmed using standard PCR conditions. PCR fragments were sequence-verified either by Sanger sequencing (*PCDH15*) or by long-read amplicon sequencing (*USH2A*). PCR conditions are available upon request.

### Variant classification

SNVs were classified according to the American College of Medical Genetics (ACMG) guidelines using Franklin (https://franklin.genoox.com/clinical-db/home), based on the automatically ran classification engine, together with manual review and SVs were manually classified according to published ACMG guidelines^31,32^. SNVs with an effect on pre-mRNA splicing were classified based on their effect in the minigene splice assay or targeted long-read cDNA transcript analysis. Variants with a full stop gain effect were classified as pathogenic, variants resulting in an in-frame effect, were classified as likely pathogenic. Probands with two (likely) pathogenic variants were considered genetically resolved, probands with one (likely) pathogenic variant and one VUS were considered possibly resolved, probands with deaf-blindness and (likely) pathogenic variants in a gene partially explaining their phenotype were considered partially resolved. Segregation analysis was performed when DNA samples from relatives were available.

### Primer sequences

All primers sequences are provided in **Supplementary table 2.**

## Results

### Composition of the study cohort

The 148 genetically unresolved probands selected for this study represent the broad clinical spectrum of IRDs (**Table 1**). The most frequently reported phenotype in the cohort was retinitis pigmentosa (53%, OMIM: 268000), followed by Usher syndrome (13%, OMIM: 276901), cone-rod dystrophy (7%, OMIM: 601777) and macular dystrophy (9%, OMIM: 616152). Other rare phenotypic subtypes, each accounting 3% or less in the cohort, included, amongst other phenotypes, Stargardt disease (OMIM: 248200), Bardet-Biedl syndrome (OMIM: 209900) and Leber congenital amaurosis (OMIM: 204000).

Following the WGS studies by Fadaie et al. and Reurink et al., hereafter referred to as “WGS studies”, a total of 127 cases remained genetically unresolved^14,16^. Four cases were resolved through earlier performed reanalysis efforts^28,33–35^. In the present study, out of the 123 unresolved cases we selected 120 cases (Fadaie et al. n=75 or Reurink et al. n=45) that were included for a stepwise, case-by-case reanalysis. Three unresolved cases were excluded from the present reanalysis effort due to a mismatch in phenotype and scope of this study. Based on availability of additional unresolved IRD samples, the analysis was expanded with the addition of these 28 cases (**Figure 1**). Contrasting the unresolved cases from the WGS studies, these additional cases had not undergone prior WGS but underwent ES or targeted single-molecule molecular inversion probe (smMIPs) sequencing for MD-, RP- or LCA-associated genes as pre-screening. An overview of the composition of the cohort is provided in **Supplementary table 1**.

**Figure 1.**
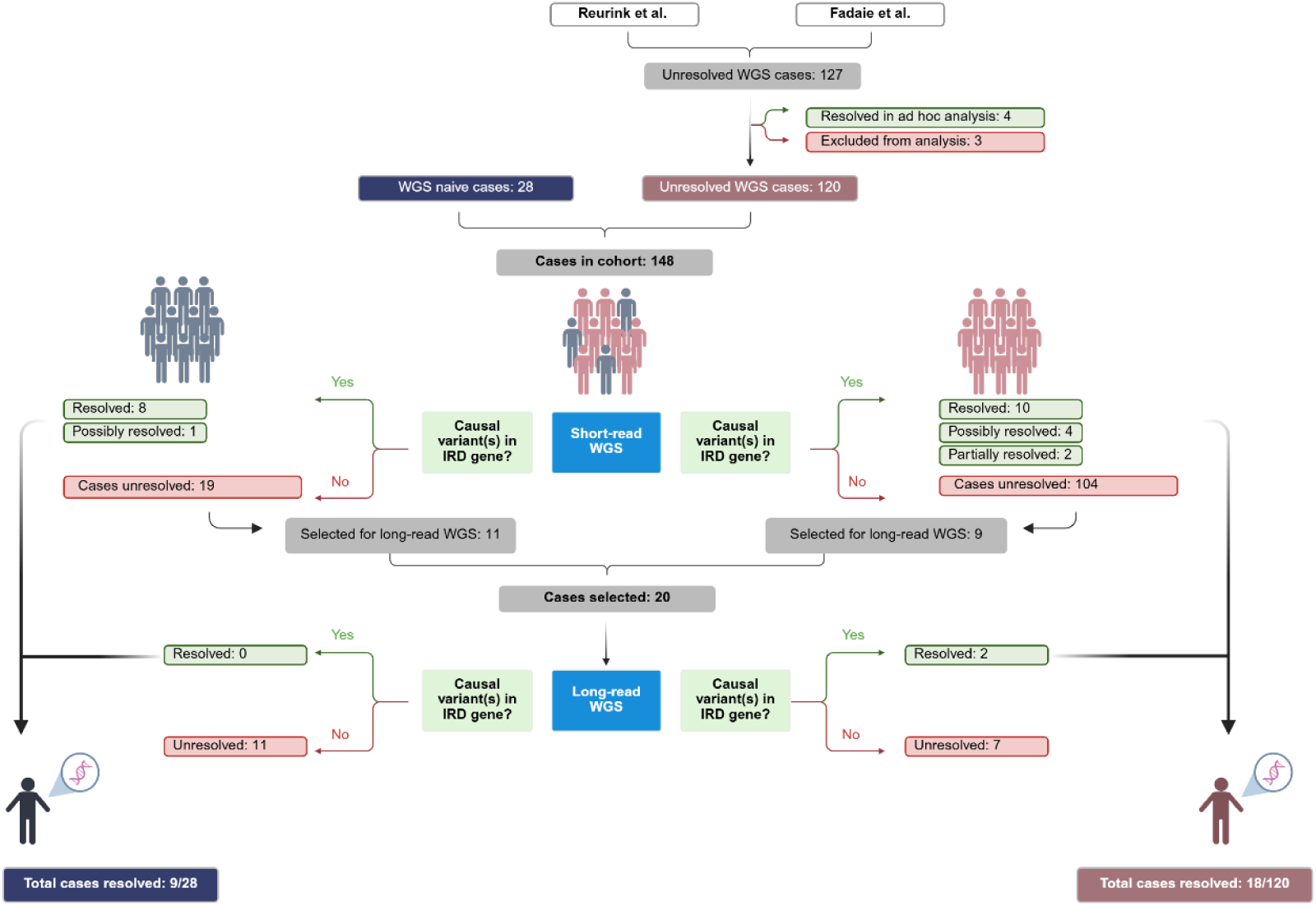
Flowchart summarizes the composition of the study cohort. A total of 127 cases remained unresolved following the initial WGS efforts by Reurink et al. and Fadaie et al. Four cases were resolved through prior ad hoc reanalysis efforts. Three unresolved cases were excluded from the present study. 120 unresolved WGS cases were combined with an additional 28 WGS naive cases, resulting in a total cohort of 148 probands. Through short-read WGS (re)analysis, cases were either resolved, possibly resolved, partially resolved, or remained unresolved. A subset of unresolved cases was selected for subsequent long-read WGS analysis which either resulted in a resolved or unresolved genetic diagnoses. Abbreviations: WGS, whole genome sequencing; IRD, inherited retinal disease

### Refined short-read genome reanalysis yields added diagnostic rate

First, all 148 probands were subjected to case-by-case (re)analysis using the refined short-read WGS workflow. In total, pathogenic or likely pathogenic variants supporting a (possible or partial) genetic diagnosis were identified in 27 of 148 cases. This included 16 diagnoses among the cases from the WGS studies cohort following short-read WGS (13%, 16/120), and nine (possible) genetic diagnoses among the 28 cases without prior WGS analysis (32%, 9/28) **(Figure 1)**. An overview of these variants is provided in **Table 2** and in the extended version of the table in **Supplementary table 3.**

**Table 2:**
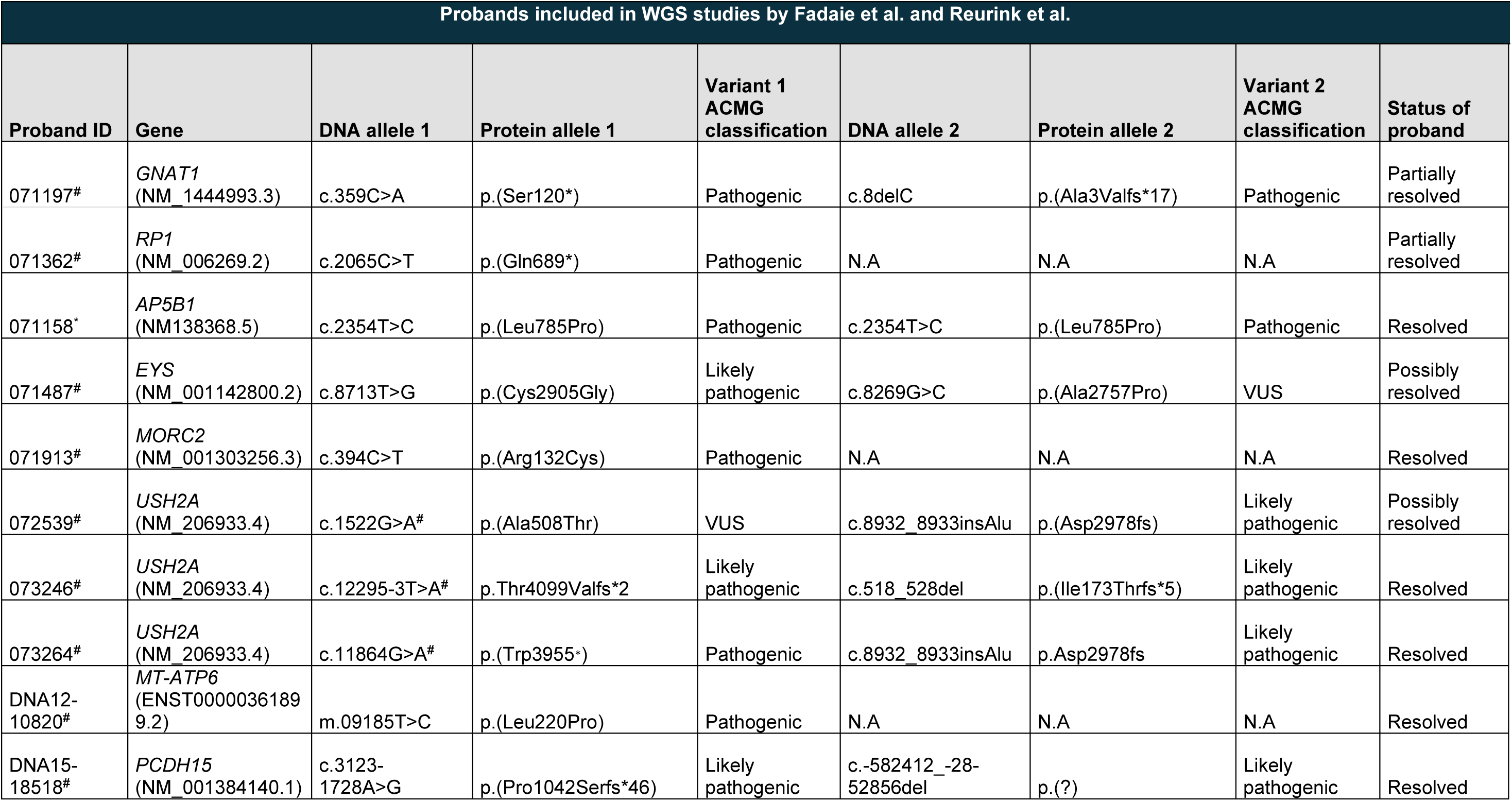
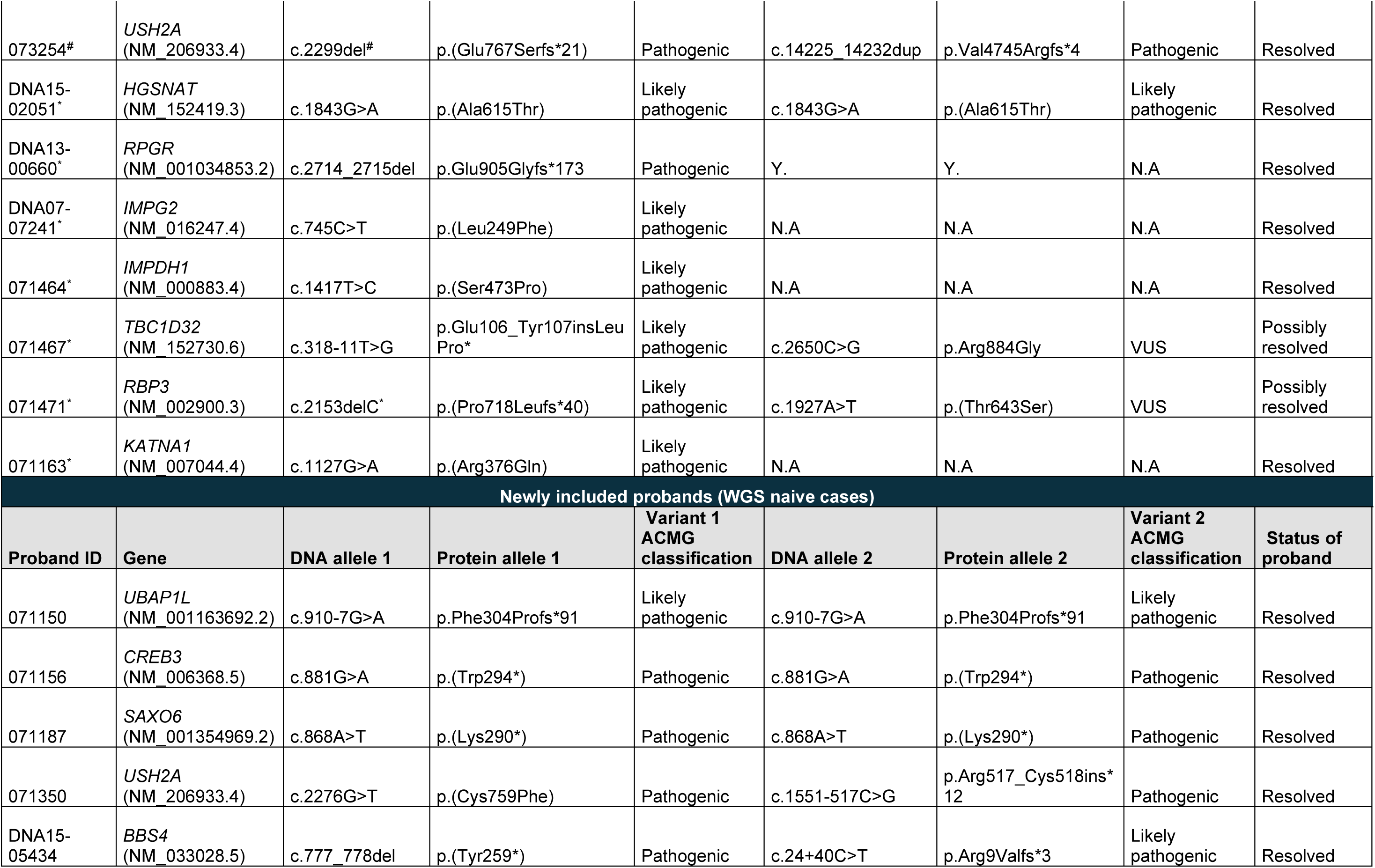
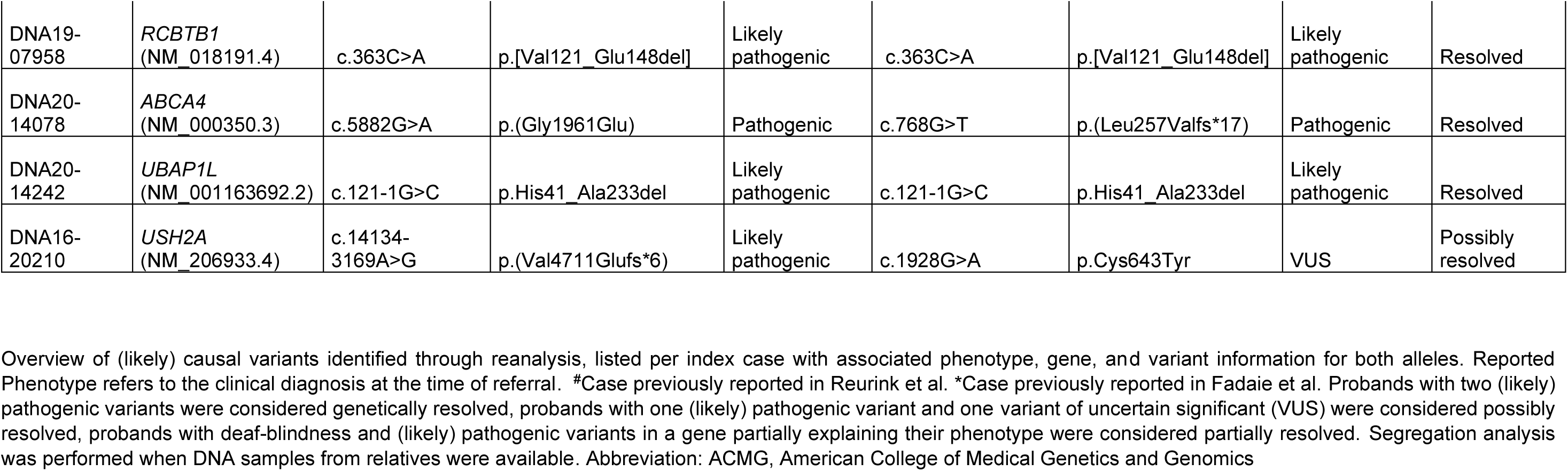
overview of (likely) causal variants identified in this study.

Five of the (likely) resolved cases had previously been reported with a monoallelic pathogenic or likely pathogenic variant in earlier WGS studies but were considered genetically unresolved, reanalysis now identified the second (likely) pathogenic allele in these individuals.

Below, the three main aspects contributing to the improved diagnostic yield are expanded on. An overview of justifications why variants were previously overlooked is provided in **Supplementary table 3.**

Overview of (likely) causal variants identified through reanalysis, listed per index case with associated phenotype, gene, and variant information for both alleles. Reported Phenotype refers to the clinical diagnosis at the time of referral. ^#^Case previously reported in Reurink et al. *Case previously reported in Fadaie et al. Probands with two (likely) pathogenic variants were considered genetically resolved, probands with one (likely) pathogenic variant and one variant of uncertain significant (VUS) were considered possibly resolved, probands with deaf-blindness and (likely) pathogenic variants in a gene partially explaining their phenotype were considered partially resolved. Segregation analysis was performed when DNA samples from relatives were available. Abbreviation: ACMG, American College of Medical Genetics and Genomics

### Variants in novel disease-associated genes contribute a quarter of the added diagnostic yield

In seven cases for whom pathogenic or likely pathogenic variants were identified, the variant was located in a gene for which the disease-gene association had only recently been established. This accounts for approximately a quarter of the total added diagnostic yield (25%, 7/27) The pathogenic variants were reported in genes (*AP5B1*, *CREB3, KATNA1, SAXO6, TBC1D32, UBAP1L*) that have only been associated with IRDs within the past years^36–41^. These novel associations are each described in separate publications, and we refer to these publications for full genetic and molecular evidence. Several of the published manuscripts directly include cases from this cohort (*AP5B1*, *CREB3, KATNA1*, *SAXO6*, *UBAP1L*) whereas the genetic cause in *TBC1D32* was identified upon publication of the novel disease-gene association.

### Splice-altering variants are a main contributor of added diagnostic yield

From the identified variants in this study, nine variants were predicted to alter pre-mRNA splicing, of which three variants are within two recently associated disease genes (*TBC1D32*: c.318-11T>G*, UBAP1L*: c.910-7G>A and c.121-1G>C) **(Table 2, Supplementary table 3**) described in the previous paragraph. As their novel disease-gene association is the primary reason these variants were previously not identified, these variants are not described in more detail here. Furthermore, a deleterious effect on gene splicing previously was demonstrated for two of the variants identified *(PCDH15* c.3123-1728A>G*, USH2A:* c.14134-3169A>G^42,43^).

To evaluate the predicted effect on pre-mRNA splicing of the four novel variants, we performed functional assays using *in vitro* minigene splice assays (*RCBTB1*, *TBC1D32*, *USH2A*) (**Figure 2A**) or targeted long-read cDNA transcript analysis (*BBS4*) using blood-derived RNA from the respective proband (**Figure 2B**). An overview of putative splice variants and SpliceAI predictions is provided in **Supplementary table 4.**

**Figure 2.**
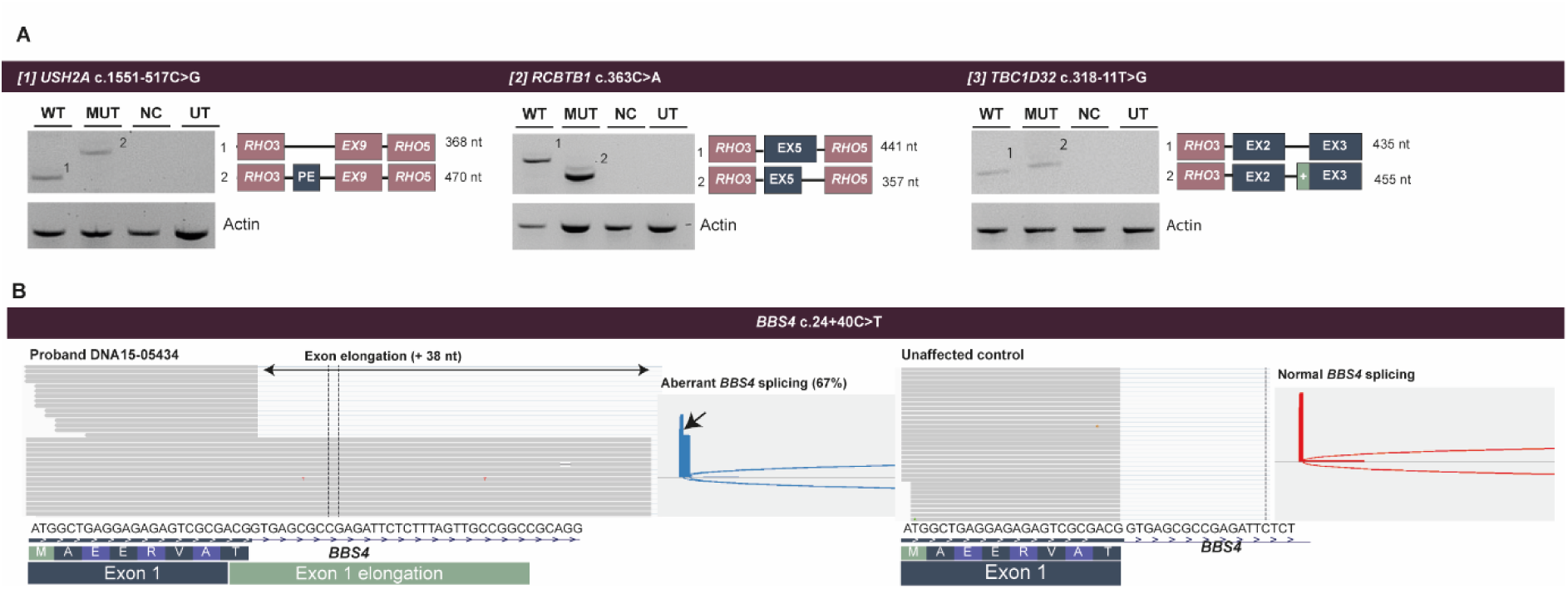
Transcript and splicing analysis of pathogenic variants. **(A)** Results of minigene splice assays for *USH2A* (c.1651-517G>C), *RCBTB1* (c.363C>A), and *TBC1D32* (c.318-11T>G) showing aberrant transcript formation compared with wildtype (WT), mutant (MUT), negative control (NC), and untransfected (UT) samples. Actin serves as a loading control. The schematic exon maps below each gel indicate the altered transcript structures and amplicon sizes**. (B)** Transcript analysis of blood-derived RNA using amplicon-based long-read sequencing confirmed the 38-nt elongation of exon 1 in 67% (19951/29679 reads) of *BBS4* transcripts (p.Arg9Valfs*43), phasing of the reads was not possible. The proband (DNA15-05434) shows aberrant *BBS4* splicing compared with unaffected control with a normal *BBS4* splicing pattern.

Deep-intronic variant *USH2A* c.1551-517C>G was predicted to induce pseudo-exon inclusion. We confirmed that the *USH2A* c.1551-517C>G variant causes the inclusion of a 102 nt pseudo-exon in intron 8 of the *USH2A* transcript, introducing a stop codon and leading to premature termination of *USH2A* protein translation (p.Arg517_Cys518ins*12). For this construct, we additionally observed skipping of in-frame exon 8 both in the control and mutant sample. Since the event was observed in both constructs, it likely reflected the assay context rather than an effect of the variant. Synonymous variant *RCBTB1* c.363C>A was predicted to result in partial exon skipping. Validation of this variant through *in vitro* minigene splice assay and subsequent sequence validation through Sanger sequencing demonstrated that *RCBTB1* c.363C>A leads to in-frame skipping of 84 nt of exon 5 (p.(Val121_Glu148del)), which encodes a part of the regulator of chromosome condensation 1 (RCC1) domain.

Two intronic variants were predicted to lead to exon-elongation. We confirmed that *TBC1D32* c.318-11T>G disrupts the polypyrimidine tract, leading to a loss of canonical 3’-splice site recognition. Through Sanger sequencing we validated the 18 nt exon-elongation resulting from the activation of an upstream cryptic splice acceptor site (p.(Glu106_Tyr107insLeuPro*)). Due to the genomic position of the splice-altering variant *BBS4* c.24+40C>T in intron 1 of the gene, the effect of this variant on pre-mRNA splicing could not be evaluated using a minigene splice assay. However, because patient-derived RNA was available, the splicing effect could be assessed directly at the endogenous transcript level, providing a more physiologically relevant evaluation. Transcript analysis of blood-derived RNA using amplicon-based long-read sequencing confirmed elongation of exon 1 by 38 nt in 67% (19951/29679 reads) of *BBS4* transcripts (p.(Arg9Valfs*43)).

### Pathogenic Alu element insertion in *USH2A* identified in two Usher syndrome cases

We assessed our cohort for the insertion of retrotransposons in IRD-associated genes using MELT. The analysis revealed an identical insertion of an Alu element in exon 45 of *USH2A* in two unrelated Usher syndrome cases.

To validate this event, we amplified the breakpoints resulting from the MEI and sequenced the inserted fragment using long-read amplicon sequencing. We confirmed the insertion of a 336 bp Alu element (*USH2A*: c.8932_8933insAlu, p.(Asp2978Glufs*75)) in both cases. The Alu insertion is present in a heterozygous state and occurs potentially *in trans* with a pathogenic variant in *USH2A* (072539 c.1522G>A, p.(Ala508Thr), 073264: c.11864G>A, p.(Trp3955*)), however, this could not be confirmed due to lack of parental DNA. The same Alu insertion was previously identified and reported as pathogenic by Torene et al.^44^.

### Long-read whole genome sequencing increases diagnostic yield through improved variant detection

From the 148 probands included in this study, 123 remained without a genetic diagnosis after our reanalysis effort. To further increase the diagnostic yield in this cohort, we performed long-read WGS for 20 cases that remained without a diagnosis. These cases were selected based on the availability of sufficient high-quality DNA material. We applied long-read WGS to address limitations of short-read WGS by improving detection of complex structural variants and to enable variant detection in genomic regions that are poorly captured by short-read WGS. Of the 20 cases, long-read WGS detected two likely pathogenic variants in two cases supporting a conclusive genetic diagnosis in two out of the 20 cases subjected to long-read WGS.

In DNA15-18518 a heterozygous deletion of 0.67 Mb was identified overlapping with the 5’-UTR and upstream region of *PCDH15*. The deletion affects the first non-coding exon and previously described *cis*-regulatory elements of the gene^45^. We performed breakpoint PCR and sequence-verified the resulting PCR fragment through Sanger sequencing. The deletion, *PCDH15*: c.-582412_-28-52856del, was confirmed in the proband, as well as in the affected sibling **(Figure 3A) (Supplementary Figure 1)**. Through segregation analysis, this variant was confirmed to be *in trans* to a second likely pathogenic variant (*PCDH15*: c.3123-1728A>G^43^) In a male proband (DNA13-00660), long-read WGS detected a known pathogenic frameshift deletion in *RPGR*, c.2714_2715del (p.(Glu905Glyfs*173)), which was not detected by short-read WGS. The variant is located within the highly repetitive ORF15 (exon 15) region of the gene **(Figure 3B)**.

**Figure 3.**
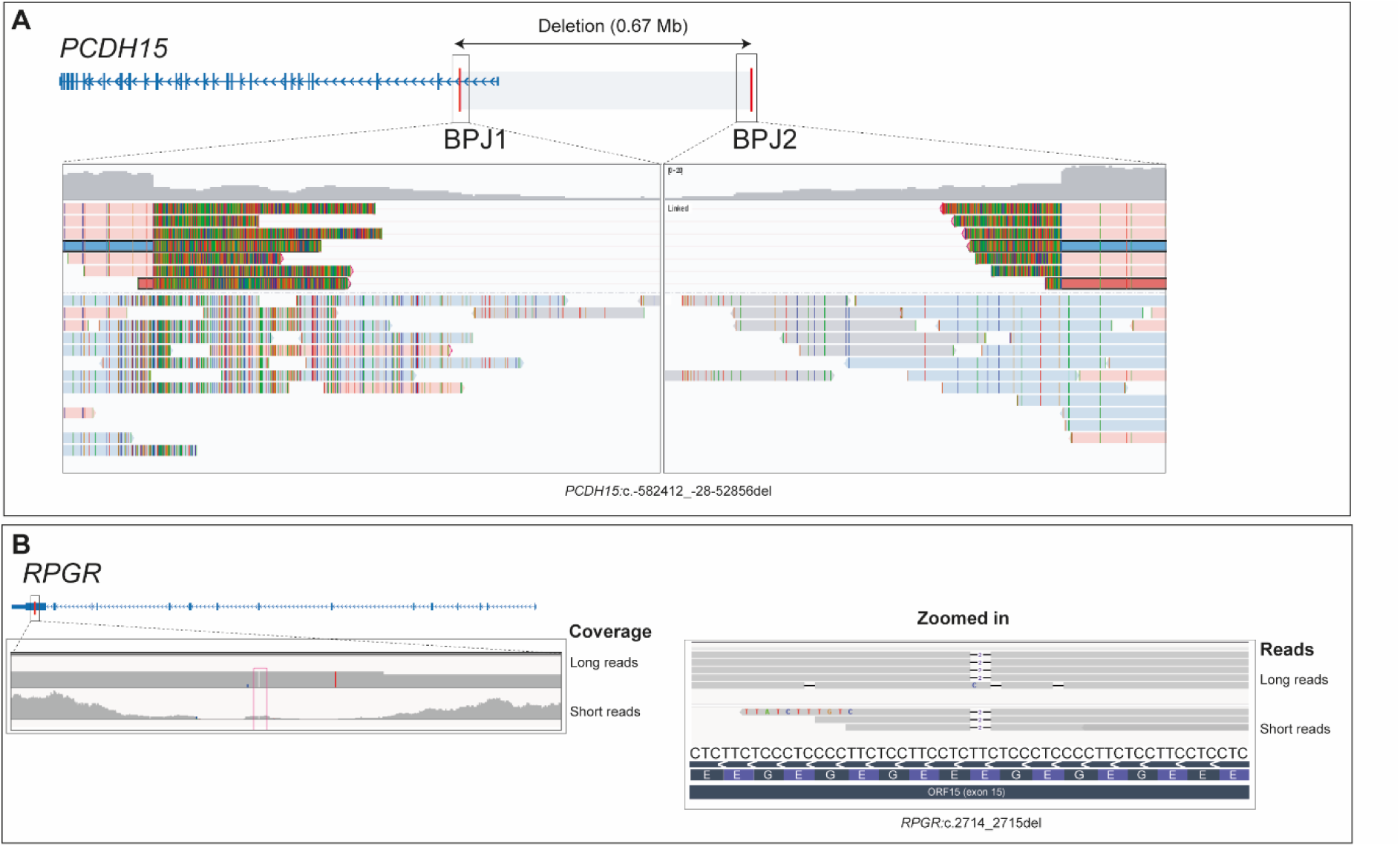
Identification of a large PCDH15 deletion and an ORF15 frameshift variant in RPGR using long-read sequencing. ***(A) PCDH15* deletion**. The *PCDH15* gene structure with exons (blue) and the 0.67 Mb deleted interval are displayed in gray. Breakpoint junctions (BPJ1 and BPJ2) are shown in the zoomed views below, where long-read alignments demonstrate reduced coverage across the deleted region and split-reads spanning the breakpoint junctions. Long-read alignments (colored bars) provide direct support for the deletion (*PCDH15*: c.-582412_-28-52856del). **(B) Frameshift variant in *RPGR* ORF15.** left panel displays short- and long-read coverage across ORF15, with long-read alignments supporting a two-base deletion (zoomed in view). The right panel shows the zoomed sequence view and highlights the affected bases.

## Discussion

Short-read WGS enables detection of nearly all classes of genomic variation and is a powerful approach for identifying the genetic causes of IRDs. We evaluated 148 probands with diverse IRD phenotypes using a case-by-case, stepwise framework that combined an updated RetNet gene list and additional filters to prioritize variants (gnomAD version 3.1.2, MELT, REVEL) with targeted functional assays. For 20 cases who remained unresolved after short-read WGS and had availability of high-quality DNA, long-read WGS was applied to capture variants potentially missed by short-read WGS. Our stepwise, case-by-case reanalysis approach of the 120 unresolved cases from the previous WGS studies yielded a solve rate of 15% (n=18 resolved (two partially resolved, four possibly resolved, 12 resolved), n=102 unresolved). Of these 18 resolved cases, long-read WGS contributed two genetic diagnosis (one structural variant and one *RPGR* ORF15 variant), demonstrating the complementary value of long reads for variant types and genomic regions that are difficult to resolve with short-read data.

Of the 28 cases without prior WGS, the stepped WGS analysis in this study resulted in an added diagnostic yield of 32% (n=9 resolved (eight resolved, one possibly resolved), n=19 unresolved).

The majority of the newly identified (likely) pathogenic variants in this research cohort were discovered through systematic reanalysis of existing short-read WGS data, emphasizing that a periodic reanalysis is a high-value, cost-effective strategy to increase the diagnostic yield in a research setting. Many previously unresolved cases did not reflect intrinsic limitations of short-read WGS but rather resulted from earlier missed variant identification and prioritization choices.

A substantial fraction of the added diagnostic yield reflects two closely related events, first the expansion of disease-gene knowledge since the original analysis, and second, the use of restricted analysis that excluded relevant causal variants. Here, we performed reanalysis of WGS data using an updated gene list, comprising all genes included in RetNet (accessed July 2026) as well as recently published IRD-associated genes. We identified variants in six genes with novel disease associations, which contributed an important part of the added diagnostic yield.

The prior use of restricted phenotype- or inheritance-based panels (restricted to genes known to be causal for autosomal recessive retinitis pigmentosa and Usher syndrome) led to two missed partial genetic diagnosis. Cases 071362 and 071197 were both reported to present with Usher syndrome, however, in both cases we identified variants to be the likely cause of RP. In individual 071362 we identified a pathogenic truncating *RP1* variant which is associated with autosomal dominant RP. In case 071197 we identified biallelic pathogenic *GNAT1* variants which are causal for congenital stationary night blindness. While the genetic analysis of genes associated with non-syndromic deafness was beyond the scope of this reanalysis effort, we expect that these causal variants are likely to be identified upon further genetic analysis and hypothesize that a dual genetic condition (autosomal dominant RP and non-syndromic hearing loss) underlies the reported phenotype in these two individuals. This illustrates how strict filtering criteria can overlook the co-existence of RP and hearing impairment as two independent genetic conditions rather than a dual genetic condition.

Population database limitations also contributed to the added diagnostic yield. A known hypomorphic *HGSNAT* founder allele (c.1843G>A, p.Ala615Thr) was excluded under a ≤0.01 cutoff in GoNL (reported frequency 1.004%)^46^. This variant was homozygous in case DNA15-02051 and supports the phenotype. The integration of larger allele frequency databases, such as gnomAD v3.1.2 aids to mitigate such exclusions.

Another major driver of the added diagnostic yield was attributed to refined variant prioritization and interpretation. SpliceAI was used as the primary tool for splice-effect prediction and we implemented a stricter, consensus-based threshold (two of four Δ scores ≥ 0.20) compared with thresholds used in earlier WGS studies (Reurink et al.: two of four Δ scores ≥ 0.10 or one Δ ≥ 0.15; Fadaie et al: at least one score Δ ≥ 0.02). While the filtering in this study is was more stringent, this reduced low-confidence candidates and focused follow-up on variants with higher predicted splice impact. Functional follow-up analysis was performed for novel splice-variants of which all the tested variants showed splice alterations consistent with SpliceAI predictions.

Subsequent long-read WGS was performed for a selected 20 cases that remained genetically unresolved following short-read WGS (re)analysis. Long-read WGS provided a genetic diagnosis for two probands. We detected a known pathogenic frameshift deletion in *RPGR*, c.2714_2715del, p.(Glu905Glyfs*173), within the highly repetitive ORF15 (exon 15) region, known to be a challenging region for sequencing with typical poor coverage in short-read WGS^47^. Retrospective inspection of short-read WGS reads confirmed very limited coverage of this region, explaining why this variant was missed using short-read WGS.

The diagnostic yield from long-read WGS (10%) is in line with several recent reports that demonstrated 7.3-11.8% additional diagnostic yield from long-read WGS^48,49^. Despite the modest overall added diagnostic yield, long-read WGS offers prominent advantages, such as phasing of compound heterozygous variants, resolving variants within highly repetitive regions, and its sensitivity to detect complex rearrangements renders the technique a valuable tool to increase the diagnostic yield in a pre-screened cohort.

Despite the comprehensive stepwise, case-by-case reanalysis, 81% of the cases (121/148) remain without a conclusive genetic diagnosis. Several factors likely contribute to missing heritability. First, our understanding and annotation of non-coding regions remains incomplete, as evidenced by the recent association of variants in the snRNA *RNU4*-2 and *RNU6* genes with autosomal dominant retinitis pigmentosa^50^. Second, in the same study, it was shown that a higher-than-expected proportion of *de novo* variants were observed in isolated cases. Only a single *de novo* variant was identified in this study (071913), most likely reflected by the limited availability of parental DNA samples. We therefore argue that *de novo* events may contribute more than anticipated to our unresolved cohort, and that trio-based sequencing should be considered when feasible.

In conclusion, our systematic periodic reanalysis of existing short-read WGS data followed by the selective long-read WGS increased the diagnostic yield substantially. Our optimized approach contributed an added diagnostic yield of 15% and 32% for the two research cohorts included in this study. Based on our findings, we recommend the following strategies for effective reanalysis of existing short-read WGS data. Existing short-read WGS data should be reanalyzed periodically (every 2-4 years) using an updated gene list, appropriate population resources and state-of-the art variant effect predictor tools. This is essential to identify variants in genes that are newly associated with IRDs. Where feasible, trio-based sequencing should be considered to increase the detection of *de novo* variants and potentially identify novel gene-disease associations. Long-read WGS should be considered for cases that remain without a conclusive genetic diagnosis to identify variants which are typically poorly captured by short-read WGS.

Although many cases of the presented research cohorts remain unresolved, our results demonstrate that a stepwise, case-by-case reanalysis followed by targeted long-read WGS substantially reduces the diagnostic gap in IRDs within a research setting. Continued improvements in the annotation of non-coding regions, broader adoption of trio sequencing, and higher-coverage long-read sequencing platforms, will be important to further close the remaining gap.

## Data availability statement

Data are available upon reasonable request. All other WGS data are subject to controlled access because they may compromise the privacy of research participants. These data may become available upon a data transfer agreement approved by the local ethics committee and can be obtained after contacting the corresponding author (S.R.) upon request.

## Acknowledgements

We thank all patients and their family members for their help and participation in this study. We thank S. van der Velde-Visser, E. Blokland and M. Jacobs-Camps for sample registration and administration. We thank the Department of Human Genetics and the Radboud Genome Technology center for infrastructural and computational support. In memoriam: Prof. Dr. Arthur A. Bergen, a valued co-author who passed away prior to the publication of this work. This work was generated within the European Reference Network for Rare Eye Diseases (ERN-EYE).

## Supplementary figures

**Supplementary figure 1.**
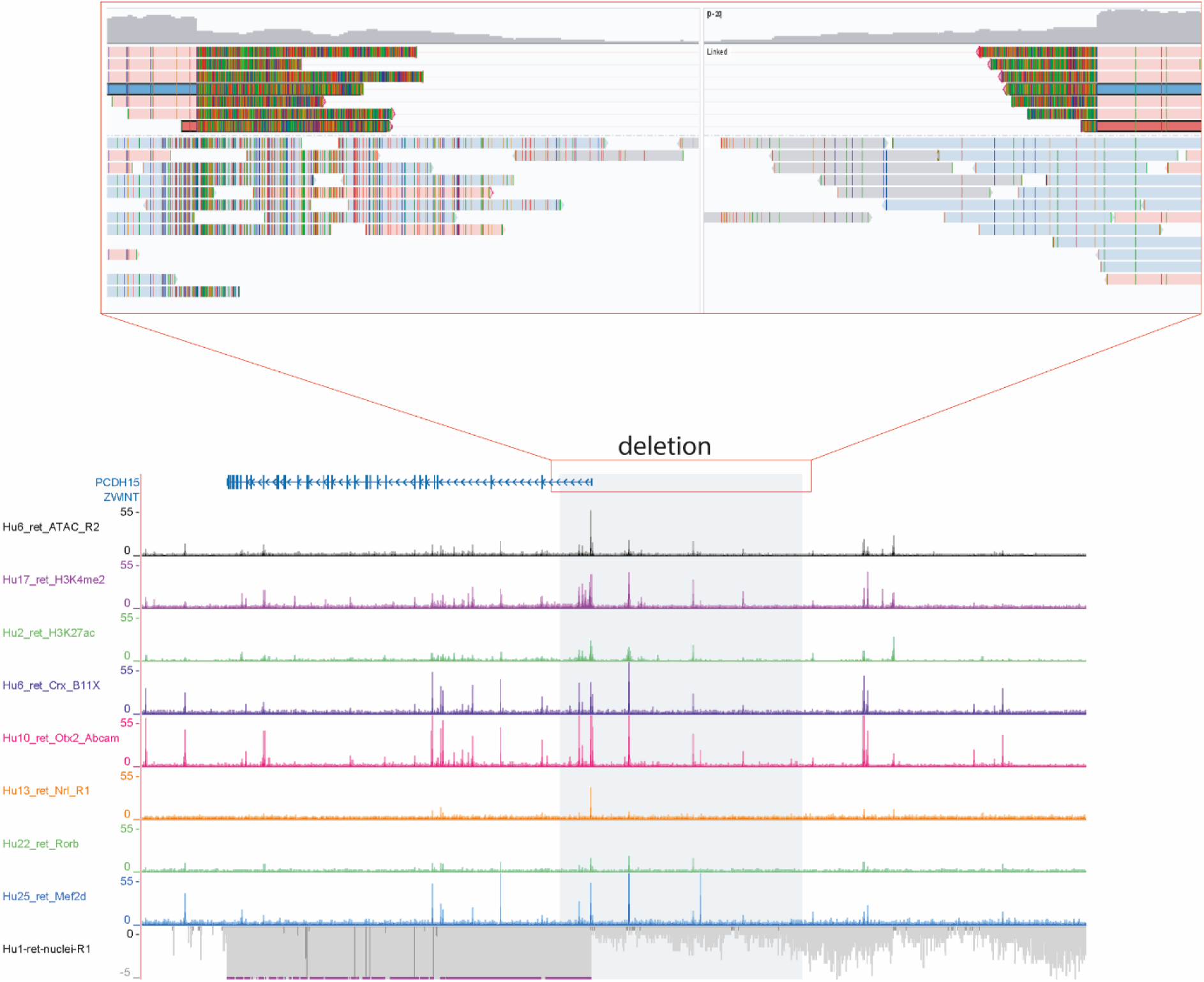
Deletion at the PCDH15 locus and its genomic regulatory context. In the lower panel, the *PCDH15* gene is displayed in blue and the 0.67 Mb deleted interval is displayed in gray, overlapping with the first non-coding exon and cis-regulatory elements of the gene. In the zoomed view above, long-read alignments demonstrate reduced coverage across the deleted region and split-reads spanning the breakpoint junctions. Long-read alignments (colored bars) provide direct support for the deletion (*PCDH15*: c.-582412_-28-52856del).

## Supplementary tables

All supplementary tables are available at: [dropbox supplementary tables Chapter 3](https://www.dropbox.com/scl/fo/ihan05j4gh3c815ub6eqh/AD06PYAmml_GcbyVeJz_G7g?rlkey=q0vee3snf07z4sg0mel08b5z6&st=34p9dd9m&dl=0)

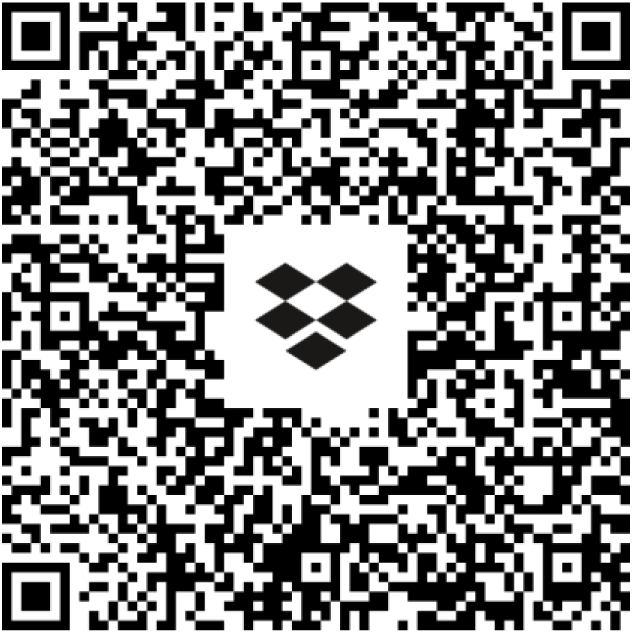

**Supplementary table 1.** overview of all cases included in the study.

| Original WGS study | Study and case ID | Phenotype | Status |
| --- | --- | --- | --- |
| Fadaie et al. | Pt-25 (071480) | STGD1 | Unresolved |
| Fadaie et al. | Pt-26(071163) | MD | Resolved |
| Fadaie et al. | Pt-27 (070250) | STGD1 | Unresolved |
| Fadaie et al. | Pt-28 (071158) | RP | Resolved |
| Fadaie et al. | Pt-29 (DNA17-02249) | RCD | Unresolved |
| Fadaie et al. | Pt-30 (DNA07-07241) | CACD | Resolved |
| Fadaie et al. | Pt-31 (DNA10-04248) | RP | Unresolved |
| Fadaie et al. | Pt-32 (071167) | Nystagmus | Unresolved |
| Fadaie et al. | Pt-33 (070256) | RP | Unresolved |
| Fadaie et al. | Pt-34 (DNA11-20872) | RP | Unresolved |
| Fadaie et al. | Pt-35 (070252) | RP | Unresolved |
| Fadaie et al. | Pt-36 (DNA08-00524) | MD | Unresolved |
| Fadaie et al. | Pt-37 (DNA13-13026) | MD | Unresolved |
| Fadaie et al. | Pt-38 ( 071179) | RP | Unresolved |
| Fadaie et al. | Pt-39 (DNA13-16873) | CD | Unresolved |
| Fadaie et al. | Pt-40 (DNA14-29307) | CRD | Unresolved |
| Fadaie et al. | Pt-41 (DNA14-25269) | MD | Unresolved |
| Fadaie et al. | Pt-42 (DNA13-02269) | RP | Unresolved |
| Fadaie et al. | Pt-43 (DNA16-08254) | CRD | Unresolved |
| Fadaie et al. | Pt-44 (DNA13-00660) | RP | Resolved |
| Fadaie et al. | Pt-45 (070265) | RP | Unresolved |
| Fadaie et al. | Pt-46 (DNA15-02051) | RCD | Resolved |
| Fadaie et al. | Pt-47 (071166) | RP | Unresolved |
| Fadaie et al. | Pt-48 (DNA10-16750) | MD | Unresolved |
| Fadaie et al. | Pt-49 (071479) | MD | Unresolved |
| Fadaie et al. | Pt-50 (071468) | RPED | Unresolved |
| Fadaie et al. | Pt-51 (DNA14-29088) | USH | Unresolved |
| Fadaie et al. | Pt-52 (DNA10-15937) | MD | Unresolved |
| Fadaie et al. | Pt-53 (071165) | RD | Unresolved |
| Fadaie et al. | Pt-54 (DNA05-01621) | BBS | Unresolved |
| Fadaie et al. | Pt-55 (071162) | RP | Unresolved |
| Fadaie et al. | Pt-56 (DNA14-21145) | RP | Unresolved |
| Fadaie et al. | Pt-57 (DNA15-18239) | RP | Unresolved |
| Fadaie et al. | Pt-59 (DNA14-25242) | LCA | Unresolved |
| Fadaie et al. | Pt-60 (DNA07-08842) | CRD | Unresolved |
| Fadaie et al. | Pt-61 (DNA13-00235) | RP | Unresolved |
| Fadaie et al. | Pt-62 (DNA17-08839) | RD | Unresolved |
| Fadaie et al. | Pt-63 (DNA13-06331) | CRD | Unresolved |
| Fadaie et al. | Pt-64 (071471) | RP | Possibly resolved |
| Fadaie et al. | Pt-65 (DNA15-20690) | RP | Unresolved |
| Fadaie et al. | Pt-66 (DNA12-18046) | RP | Unresolved |
| Fadaie et al. | Pt-67 (DNA13-08681) | CRD | Unresolved |
| Fadaie et al. | Pt-68 (070253) | CSNB | Unresolved |
| Fadaie et al. | Pt-69 (071464) | RP | Resolved |
| Fadaie et al. | Pt-70 (DNA14-34434) | CD | Unresolved |
| Fadaie et al. | Pt-71 (071478) | RP | Unresolved |
| Fadaie et al. | Pt-72 (071477) | CRD | Unresolved |
| Fadaie et al. | Pt-73 (071475) | RP | Unresolved |
| Fadaie et al. | Pt-74 (071474) | RP | Unresolved |
| Fadaie et al. | Pt-75 (071472) | RP | Unresolved |
| Fadaie et al. | Pt-76 (071467) | RP | Possibly resolved |
| Fadaie et al. | Pt-77 (071466) | RP | Unresolved |
| Fadaie et al. | Pt-78 (071465) | ACHM | Unresolved |
| Fadaie et al. | Pt-79 (071461) | CD | Unresolved |
| Fadaie et al. | Pt-80 (071459) | RP | Unresolved |
| Fadaie et al. | Pt-81 (070257) | RP | Unresolved |
| Fadaie et al. | Pt-82 (070249) | RP | Unresolved |
| Fadaie et al. | Pt-83 (070251) | STGD1 | Unresolved |
| Fadaie et al. | Pt-84 (070246) | STGD1 | Unresolved |
| Fadaie et al. | Pt-85 (070258) | RP | Unresolved |
| Fadaie et al. | Pt-86 (070262) | RP | Unresolved |
| Fadaie et al. | Pt-87 (070264) | RP | Unresolved |
| Fadaie et al. | Pt-88 (070259) | CRD | Unresolved |
| Fadaie et al. | Pt-89 (070260) | RP | Unresolved |
| Fadaie et al. | Pt-90 (071168) | RP | Unresolved |
| Fadaie et al. | Pt-91 (071169) | RP | Unresolved |
| Fadaie et al. | Pt-92 (071173) | FAP | Unresolved |
| Fadaie et al. | Pt-93 (071176) | RP | Unresolved |
| Fadaie et al. | Pt-94 (DNA13-16218) | MD | Unresolved |
| Fadaie et al. | Pt-95 (DNA14-23850) | RD | Unresolved |
| Fadaie et al. | Pt-96 (071177) | RP | Unresolved |
| Fadaie et al. | Pt-97 (071178) | RP | Unresolved |
| Fadaie et al. | Pt-98 (DNA13-12161) | RP | Unresolved |
| Fadaie et al. | Pt-99 (DNA13-13366) | RP | Unresolved |
| Fadaie et al. | Pt-100 (DNA15-11058) | MD | Unresolved |
| New case | 071150 | RP | Resolved |
| New case | 071156 | MD | Resolved |
| New case | 071350 | RP | Resolved |
| New case | DNA14-30406 | RP | Unresolved |
| New case | DNA15-05434 | LCA | Resolved |
| New case | DNA19-07958 | RPED | Resolved |
| New case | DNA20-14078 | STGD1 | Resolved |
| New case | DNA20-14242 | CD | Resolved |
| New case | 071132 | RP | Unresolved |
| New case | 071187 | RP | Resolved |
| New case | 074547 | USH | Unresolved |
| New case | DNA20-03286 | ACHM | Unresolved |
| New case | 071123/071124 | CRD | Unresolved |
| New case | 071127/071128 | CRD | Unresolved |
| New case | DNA21-01444 | RP | Unresolved |
| New case | DNA20-00820 | RD | Unresolved |
| New case | DNA20-00137 | RP | Unresolved |
| New case | DNA19-17346 | RP | Unresolved |
| New case | DNA19-05402 + DNA20-02672 | RP | Unresolved |
| New case | DNA19-07592 | MD | Unresolved |
| New case | DNA17-20538 | RD | Unresolved |
| New case | DNA16-20210 | RP | Possibly resolved |
| New case | DNA15-02042 | RP | Unresolved |
| New case | DNA14-26565 + DNA18-11908 | RP | Unresolved |
| New case | 74570 | RP | Unresolved |
| New case | DNA17-07703 | RP | Unresolved |
| New case | 074547 | USH | Unresolved |
| New case | DNA17-16201 | MD | Unresolved |
| Reurink et al. | 071146 | RP | Unresolved |
| Reurink et al. | 071164 | USH | Unresolved |
| Reurink et al. | 071171 | USH | Unresolved |
| Reurink et al. | 071197 | USH | Partially resolved |
| Reurink et al. | 071345 | RP | Unresolved |
| Reurink et al. | 071362 | USH | Partially resolved |
| Reurink et al. | 071365 | USH | Unresolved |
| Reurink et al. | 071485 | RP | Unresolved |
| Reurink et al. | 071487 | RP | Possibly resolved |
| Reurink et al. | 071489 | RP | Unresolved |
| Reurink et al. | 071913 | USH | Resolved |
| Reurink et al. | 071915 | USH | Unresolved |
| Reurink et al. | 071949 | USH | Unresolved |
| Reurink et al. | 071953 | USH | Unresolved |
| Reurink et al. | 071957 | RP | Unresolved |
| Reurink et al. | 071958 | RP | Unresolved |
| Reurink et al. | 071959 | RP | Unresolved |
| Reurink et al. | 072533 | RP | Unresolved |
| Reurink et al. | 072534 | RP | Unresolved |
| Reurink et al. | 072538 | RP | Unresolved |
| Reurink et al. | 072539 | USH | Possibly resolved |
| Reurink et al. | 072901 | RP | Unresolved |
| Reurink et al. | 073246 | RP | Resolved |
| Reurink et al. | 073248 | RP | Unresolved |
| Reurink et al. | 073250 | RP | Unresolved |
| Reurink et al. | 073253 | RP | Unresolved |
| Reurink et al. | 073254 | USH | Resolved |
| Reurink et al. | 073261 | RP | Unresolved |
| Reurink et al. | 073262 | RP | Unresolved |
| Reurink et al. | 073263 | RP | Unresolved |
| Reurink et al. | 073264 | USH | Resolved |
| Reurink et al. | DNA06-00907 | USH | Unresolved |
| Reurink et al. | DNA07-11651 | USH | Unresolved |
| Reurink et al. | DNA09-14634 | RP | Unresolved |
| Reurink et al. | DNA10-12537 | RP | Unresolved |
| Reurink et al. | DNA12-10820 | USH | Resolved |
| Reurink et al. | DNA13-03665 | RP | Unresolved |
| Reurink et al. | DNA15-02640 | CRD | Unresolved |
| Reurink et al. | DNA15-05363 | RP | Unresolved |
| Reurink et al. | DNA15-16748 | RP | Unresolved |
| Reurink et al. | DNA15-18518 | USH | Resolved |
| Reurink et al. | DNA17-06665 | RP | Unresolved |
| Reurink et al. | DNA17-11021 | RP | Unresolved |
| Reurink et al. | DNA17-13359 | RP | Unresolved |
| Reurink et al. | DNA18-00286 | RP | Unresolved |

Abbreviations: USH, Usher syndrome; RP, retinitis pigmentosa; HL, hearing loss; CSNB, congenital stationary night blindness; CRD, cone-rod dystrophy; CACD, central areolar choroidal dystrophy; MD, macular dystrophy; LCA, Leber congenital amaurosis; RPED, retinal pigment epithelium dystrophy; STGD1, Stargardt disease type 1; BB, Bardet-Biedle syndrome

**Supplementary table 2.**
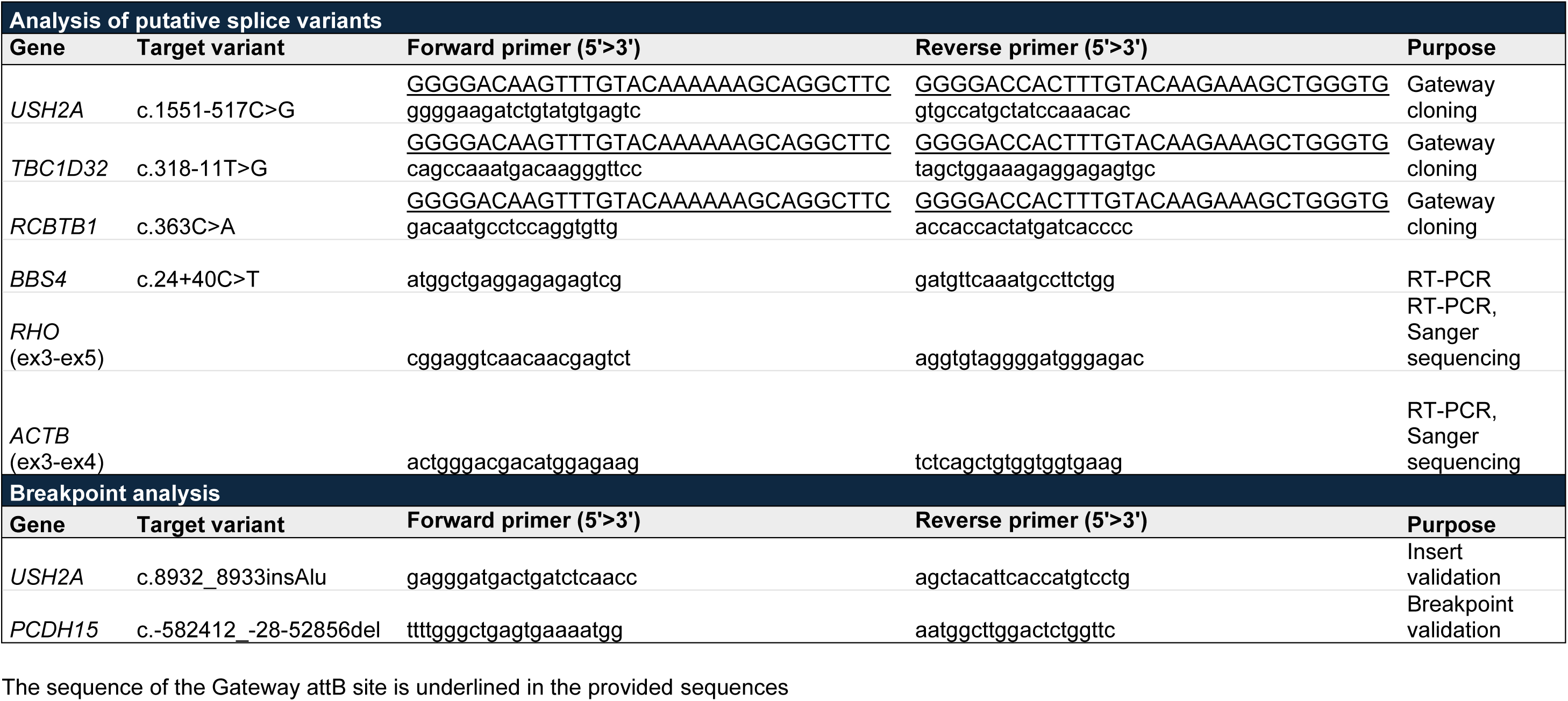
Sequences of primers used in this study The sequence of the Gateway attB site is underlined in the provided sequences.

**Supplementary table 3:** Extended overview of (likely) causal variants identified in this study

Available at: **dropbox supplementary tables Chapter 3**

**Supplementary table 4:** Overview of putative splice variants and SpliceAI predictions Splice predictions were obtained with SpliceAI and stated as gain or loss at the predicted nucleotide position from the variant. AG: accepter gain, AL: acceptor loss, DG: donor gain, DL: donor loss, Δ score: delta score. GnomAD AF: Genome Aggregation Database allele frequency in genome sequencing data. Nt: nucleotide, NA: not applicable.

| Proband ID | Gene | Variant position | SpliceAI_AG<br>Δ score | SpliceAI_AL<br>Δ score | SpliceAI_DG<br>Δ score | SpliceAI_DL<br>Δ score | Splice defect confirmed? |
| --- | --- | --- | --- | --- | --- | --- | --- |
| 071350 | USH2A | c.1551-517C>G | 0.32 (+106 nt) | n.a | 0.90 (+5 nt) | 0.04 (-11 nt) | Yes |
| 071467 | TBC1D32 | c.318-11T>G | 0.19 (+7 nt) | 0.87 (-11 nt) | n.a | n.a | Yes |
| DNA15-05434 | BBS4 | c.24+40C>T | n.a | n.a | 0.96 (-2 nt) | 0.23 (+3 nt) | Yes |
| DNA19-07958 | RCBTB1 | c.363C>A | n.a | 0.15 (+2 nt) | 0.61 (+3 nt) | 0.03 (-81 nt) | Yes |

